# Analysis of COVID-19 cases and associated ventilator requirement in Indian States

**DOI:** 10.1101/2020.07.13.20153056

**Authors:** V Dhanya, R Anitha, Ashwini Kumar Kishan, S R Sumathi, Amrit Roy

## Abstract

Analysis of COVID-19 cases and prediction of quantity of associated ventilator requirement is very relevant during this pandemic. This paper presents a method for predictive estimation of ventilator requirement for COVID-19 patients in Indian states. It uses ARIMA (Autoregressive Integrated Moving Average) model for predicting the future cumulative cases and daily fatality. Taking cue from this, ventilator requirement is estimated for each state. State wise estimation of ventilator is important because public healthcare system in India is managed at state level. Dataset on Novel Corona Disease 2019 in India from Kaggle website is used in this work.

## 1. Introduction

Coronavirus disease (COVID-19) is an infectious disease caused by coronavirus discovered in 2019.This virus is mainly transmitted through droplets generated when an infected person coughs, sneezes, or exhales. These droplets are too heavy to hang in the air, and quickly fall on floors or surfaces. People can be infected by breathing in the virus if they are within close proximity of someone who has COVID-19, or by touching a contaminated surface and then touching their eyes, nose or mouth. As per clinical management guidelines of World Health Organization (WHO), most people with COVID −19 develop mild illness (81%), and 14% develop severe disease that require hospitalisation and oxygen support and 5% require admission to an intensive care unit [1]. Most of the critically ill patient will require mechanical ventilation. Most common diagnosis in severe COVID-19 patients is severe pneumonia. In humans, several corona viruses cause respiratory infections ranging from the common cold to more severe diseases such as Middle East Respiratory Syndrome (MERS) and Severe Acute Respiratory Syndrome (SARS) [1].

In India first COVID-19 case is reported for a student who returned from Wuhan to Kerala on 30 January 2020. On 03 February 2020, India reported 3^rd^ case and there was no increase in cases till 01 March 2020. After that there was an increase in the number of reported cases. Cumulative cases in the end of June reached 5.48 lakhs. From 25 January 2020 to till date, India has taken the preventive measures such as health screening at airports and state borders, introduction of quarantine policies, visa restrictions etc. to contain the virus spread. Indian government also started five phases of lockdown from 25 March 2020 to 30 June 2020 with several restrictions such as restricting the movement of entire population of India, closure of all places of worship, all services and shops closed except pharmacies, hospitals, banks, grocery shops and other essential services etc. and relaxations given based on zone basis such as green (areas with zero confirmed cases till date or no confirmed case in the last 21 days.), red (areas or the hotspots classified as those with the highest caseload) and orange (areas which have reported a limited number of cases in the past and no surge in positive cases recently) [2].

As per press release by Press Information Bureau (PIB), on 8 May 2020, on average 4.7% of COVID-19 patient required ICU, 1.1% required ventilator support and 3.2% were on oxygen support [3]. Until 01 May 2020, India had 19,398 ventilators available for COVID-19 care and Government ordered additional 60,000 ventilators [3]. Rishabh Tyagi et. al, proposed a methodology to find the ventilators required for Indian states. In this paper ventilator requirement is calculated based on the five percent of the forecasted active cases [4]. As per worldometer, serious/ critical patient out of total case is 1.5% for India [5]. Most of the serious/critical patient require ventilators support. In this paper, a method for predicting the ventilator requirement for each Indian state by using predicted cumulative confirmed cases and daily fatality of each Indian states is presented. Estimating the ventilator requirement for all Indian states is necessary because each state has different number of confirmed cases, fatality and infection rate and also each state has public health care system managed by the state itself. And the estimation of ventilator in each state will help the local authorities in states to make the decision on ventilator requirement in future. So, the prediction of ventilator requirement is very relevant at this stage.

## 2. Data Source

Data used for this model is taken from Kaggle website [6]. This data is verified by using the daily dataset available in Ministry of Health & Family Welfare, Govt. of India website [7]. This is a time series data with date, state/union territory, cured, death and confirmed as data field. Daily fatality for each day is calculated from the cumulative death data. The data is accessed on 29 June 2020 and data used for this study is from 31 January 2020 to 29 June 2020 (151 days).

## 3. Methodology

Model selected to predict the future points of in this data is ARIMA model and it is implemented in Python language. ARIMA is a forecasting algorithm based on the idea that the information in the past values of the time series can alone be used to predict the future values. This model is represented by various parameters and model is expressed as *ARIMA (p, d, q)* [8]. Here, *p* stands for the order of auto-regression, *d* is the degree of trend difference while *q* indicates the order of moving average. This model is implemented in Python.

Parameters such as p and q for ARIMA model is arrived by plotting the Autocorrelation graph (ACF) and Partial Autocorrelation (PACF) of confirmed cases of all states. Augmented Dickey Fuller (ADF) test is used for finding the *d* value for ARIMA model [8]. If *p* value of ADF test is greater than the significance level 0.05 then the time series is not stationary. Stationarity of data is achieved by differencing of data. Initial parameters of ARIMA model is arrived by using the above method and model is evaluated based on how well it fits the data or its ability to accurately predict the future data points. Akaike Information Criterion (AIC) value is used for evaluating the model [9]. A model parameter that fits the data very well while using lots of features will be assigned a larger AIC score. Aim is to find the model with fewer features that yield lowest AIC value.

Ventilator requirement for each state is calculated by using the fatality rate.

Fatality Rate = fatality in i ^th^ day / cumulative confirmed cases

Fatality rate for the training data and predicted data is calculated and mean of that value is used for calculating the ventilator requirement in each state.

## 4. Result and Discussion

Prediction for the period 30 June 2020 to 01 August 2020 is carried out for all states in India. State which has the highest number of confirmed case (Maharashtra) is considered for discussion. Similar way other states study is carried out for the prediction. Figure 1 shows the ACF and PACF plots of Confirmed cases for Maharashtra. According to PACF and ACF plot, p and q values are selected and further tuning of the p and q parameters are carried out according to the AIC value of the model. Final selection of p and q parameters are done by using the lowest AIC values given by the model.

**Figure 1:**
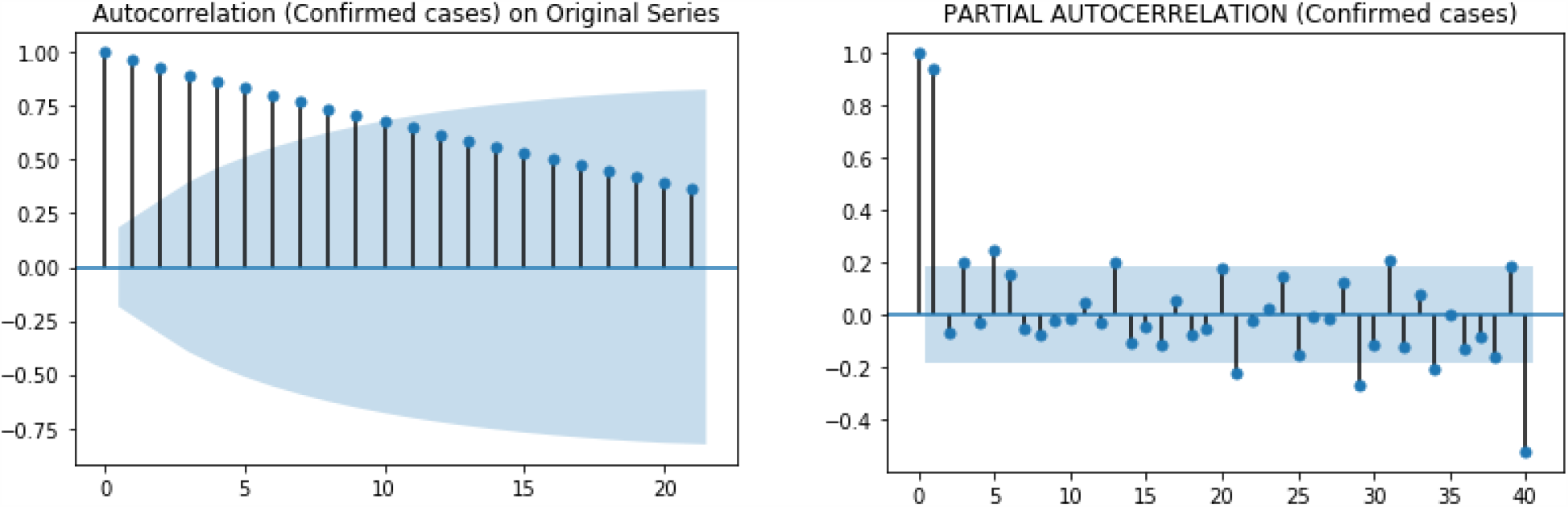
Autocorrelation and PACF plot for confirmed cases in Maharashtra

For comparing the actual and forecasted confirmed COVID-19 cases, a time series graph is plotted starting from 10 March 2020 to 29 June 2020. The plot is represented by Figure 2. Prediction plot shows the model is following the trend of the actual data. This reveals the precision of the model in forecasting.

**Figure 2:**
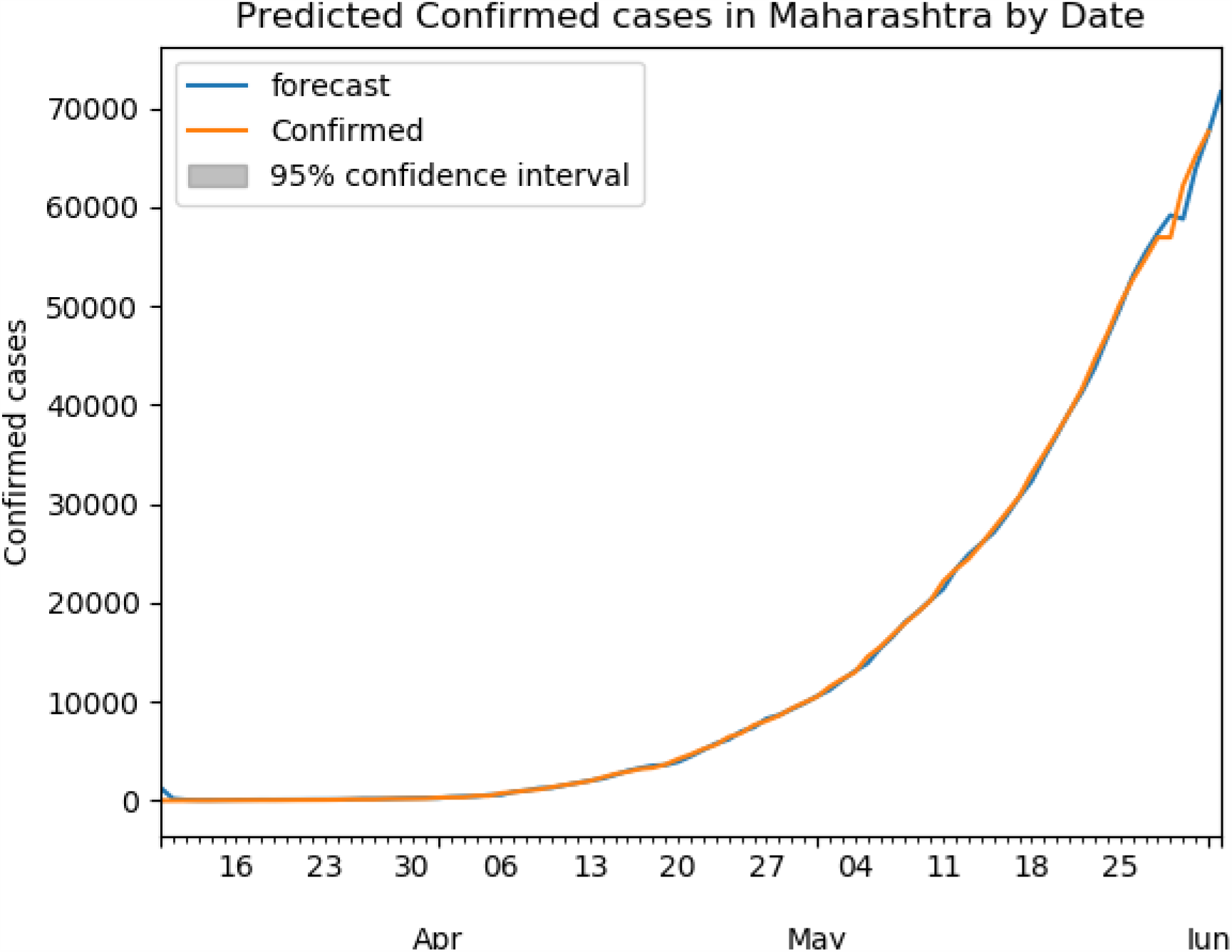
Prediction of confirmed cases from 10 March 2020 to 29 June 2020

Future prediction of confirmed cases for next 33 days from 30 June 2020 to 01 August 2020 with 95% Confidence Interval is shown in Figure 3. Number of confirmed cases are in increasing trend. This indicates that the time series data is not stationary.

**Figure 2:**
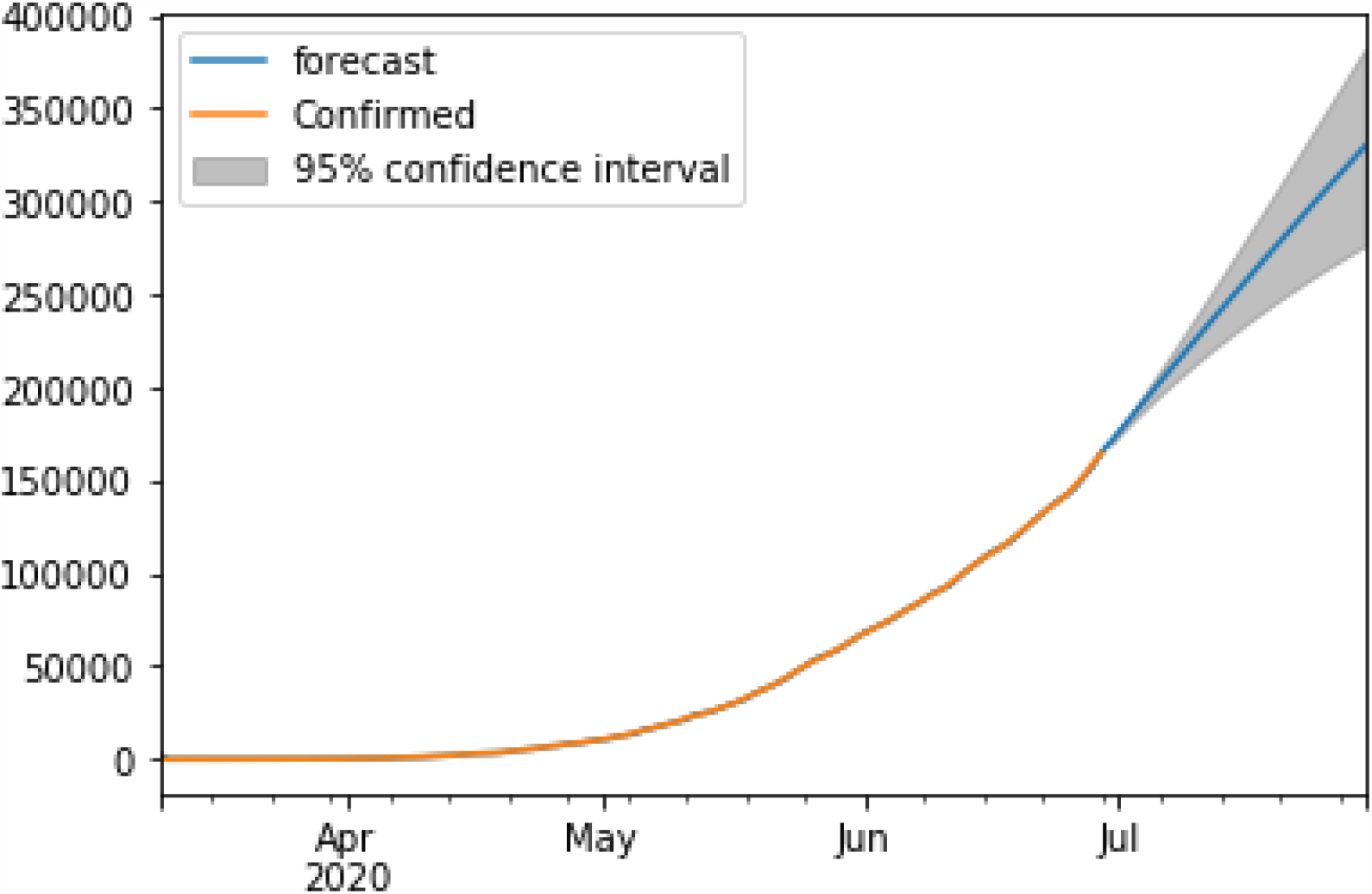
Prediction for a period from 10 March 2020 to 01 August 2020 confirmed cases in Maharashtra

Table 1 shows the predicted confirmed cases in all states and Union Territories (UT) in India. Confirmed case for next 33 days is predicted and predicted data with lower and upper limit with 95% Confidence Interval (CI) are shown in the Table 1. Most of the states and union territories are showing the increasing trend in the number of confirmed cases.

**Tab le 1:** Prediction of confirmed cases for all states and UT with 95% CI

| **States & Model with**<br>**(p, d, q)** | **02 July 2020**<br>**Predicted value**<br>**(lower limit, upper limit)** | **15 July 2020**<br>**Predicted value**<br>**(lower limit, upper limit)** | **01 August 2020**<br>**Predicted value**<br>**(lower limit, upper limit)** |
| --- | --- | --- | --- |
| Andaman and Nicobar<br>Islands (1 1 1) | 84<br>(76, 91) | 104<br>( 75 , 133 ) | 122<br>( 74, 169) |
| Andhra Pradesh<br>(1 1 0) | 15980<br>( 15425, 16535) | 25973<br>( 21015, 30931) | 35748<br>( 23693, 47804) |
| Arunachal Pradesh<br>(2 1 2) | 201<br>( 187, 215) | 272<br>( 225, 319) | 332<br>( 210, 455) |
| Assam<br>(2 1 0) | 7982<br>( 7745, 8218) | 11295<br>( 9652, 12937) | 15102<br>( 10995, 19209) |
| Bihar<br>(4 1 0) | 9871<br>( 9625, 10117) | 12630<br>( 11371, 13888) | 16017<br>( 13006 , 19028) |
| Chandigarh<br>(3 1 0) | 438<br>( 417, 459) | 488<br>( 409, 566) | 557<br>( 433, 680 ) |
| Chhattisgarh<br>(3 1 0) | 2824<br>( 2703, 2945) | 3409<br>( 2696, 4121) | 3976<br>( 2549, 5403) |
| Dadar Nagar Haveli<br>(1 1 0) | 45<br>( 24. , 67) | 55<br>( 27, 84) | 69<br>( 34, 103) |
| Delhi (1 1 0) | 91224<br>( 88869. , 93578) | 119495<br>( 99696,139293) | 145767<br>(100536,190998) |
| Goa (1 1 1) | 1375<br>( 1309 , 1441) | 2144<br>( 1774, 2515) | 3150<br>( 2198, 4103) |
| Gujarat (2 1 2) | 33084<br>( 32698, 33470) | 40787<br>( 38352, 43223) | 50910<br>( 44486, 57334) |
| Haryana (1 1 4) | 15134<br>( 14925, 15343) | 20456<br>( 18386, 22525 ) | 26844<br>( 21192, 32496) |
| Himachal Pradesh<br>(2 1 0) | 977<br>( 936 , 1019) | 1155<br>( 925, 1384) | 1326<br>( 918 , 1734 ) |
| Jammu and Kashmir<br>(3 1 0) | 7559<br>( 7318, 7800) | 9110<br>( 7621, 10599) | 10590<br>( 7536, 13645) |
| Jharkhand (2 1 0) | 2460<br>( 2345, 2575) | 2820<br>( 2169, 3471) | 3256<br>( 2073, 4438) |
| Karnataka (2 1 0) | 16923<br>( 16311, 17534 ) | 31857<br>( 25961 , 37752 ) | 48800<br>( 33202 , 64397 ) |
| Kerala (3 1 0) | 4635<br>( 4579, 4691) | 6548<br>( 6070 , 7027 ) | 9049<br>( 7702, 10396) |
| Ladakh (1 1 0) | 982<br>( 865, 1098) | 1088<br>( 709, 1467) | 1229<br>( 663, 1794) |
| Madhya Pradesh<br>(3 1 0) | 13744<br>( 13487, 14000) | 15840<br>( 14237, 17444) | 18183<br>(14942,21423) |
| Maharashtra (1 1 0) | 180860<br>(178340,183381) | 247271<br>(222743, 271799) | 325687<br>(259892,391482) |
| Manipur (1 1 0) | 1328<br>( 1223, 1432) | 1537<br>( 1165, 1908 ) | 1764<br>( 1203, 2326) |
| Meghalaya (4 1 0) | 48<br>( 42, 54) | 56<br>( 42, 70) | 66<br>( 46, 87) |
| Mizoram (2 1 0) | 152<br>( 132 , 172) | 171<br>( 103, 239) | 197<br>( 94, 299) |
| Odisha (3 1 0) | 7269<br>( 7139, 7399) | 10015<br>( 8990, 11039) | 13348<br>( 10646, 16051) |
| Puducherry (1 1 0) | 636<br>( 575 , 696 ) | 713<br>( 568 , 859 ) | 815<br>( 605 , 1025 ) |
| Punjab (1 1 0) | 5577<br>( 5328, 5826 ) | 6418<br>( 5199 , 7638) | 7258<br>( 5269, 9246) |
| Rajasthan ( 4 1 0) | 18206<br>( 17979 , 18433) | 22089<br>( 20479, 23699) | 26770<br>( 22625, 30915) |
| Tamil Nadu (1 1 0) | 94061<br>( 93082 , 95041 ) | 144571<br>( 134572 ,154571 ) | 209272<br>(180800 ,237744 ) |
| Telengana (1 1 0) | 5462<br>( 4332 , 6591 ) | 6031<br>( 4519 , 7543 ) | 6774<br>( 4874 , 8674 ) |
| Tripura (1 1 0) | 1391<br>( 1280 , 1503 ) | 1600<br>( 1295 , 1904 ) | 1873<br>( 1428 , 2318 ) |
| Uttarakhand (2 1 0) | 2952<br>( 2824 , 3081 ) | 3375<br>( 2650 , 4100 ) | 3841<br>( 2507 , 5175 ) |
| Uttar Pradesh (1 1 0) | 23461<br>( 22553 , 24370 ) | 26582<br>( 22427 , 30737 ) | 29919<br>( 23286 , 36552 ) |
| West Bengal (2 1 0) | 18921<br>( 18686 , 19156 ) | 25874<br>( 23800 , 27947 ) | 34537<br>( 28857 , 40217 ) |

Maharashtra will cross more than 325687 confirmed cases by 01 August 2020. Delhi will cross more than 145767 confirmed cases by 01 August 2020, TamilNadu will cross more than 209272 confirmed cases by 01 August 2020 and Gujarat will cross more than 50910 cases by 01 August 2020. West Bengal will cross more than 34537 cases, Uttar Pradesh will cross more than 29919 cases, Rajasthan will cross more than 26770 cases, Madhya Pradesh will cross more than 18183 cases by 01 August 2020. Goa will cross more than 3150 cases. Andaman and Nicobar Islands, Chandigarh, Dadar Nagar Haveli, Ladakh, Manipur, Meghalaya will have only less than 600 and Puducherry will be less than 1000 cases confirmed cases by 01 August 2020.

Daily fatality in each state and UT are predicted and it is shown in Table 2. Andaman and Nicobar Islands, Dadar Nagar Haveli, Ladakh, Manipur, Mizoram and Tripura are having zero daily fatality in all reported dates. ADF statistics of these states are giving not a number as the result and this data is analysed and found that mean of the data is one, variance of the data is zero and lagged difference between the values are zero. In this case, not able to find a linear regression model for fitting this value for ADF test. Because of that prediction is not with possible for these data with ARIMA model. Maharashtra will have the highest number of daily fatality compared to all states followed by Delhi, Tamil Nadu and Gujarat. Table 2 shows the predicted value and lower limit and upper limit of predicted value with 95% CI. Negative value is considered as zero for reading.

**Table 2:**
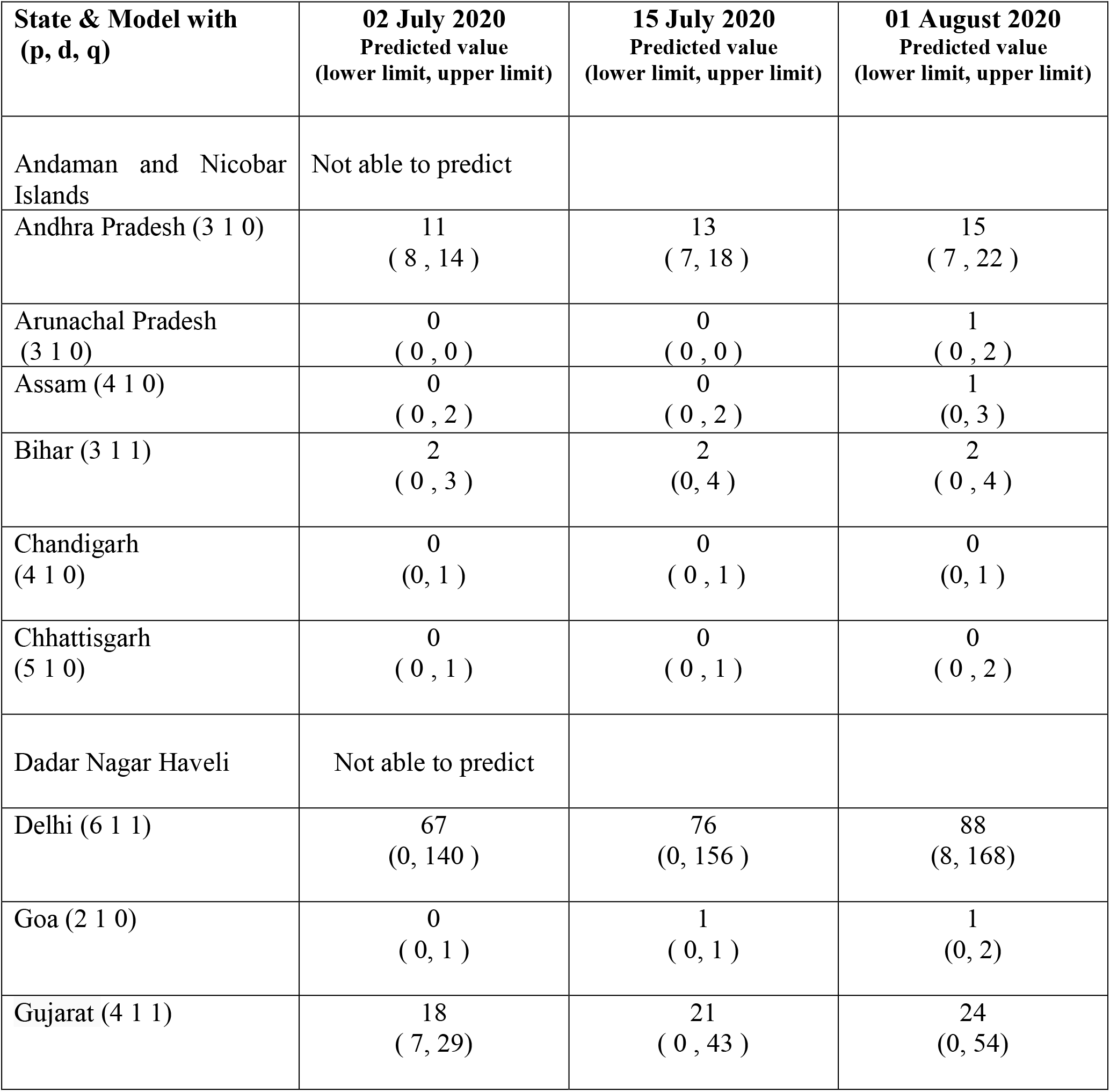
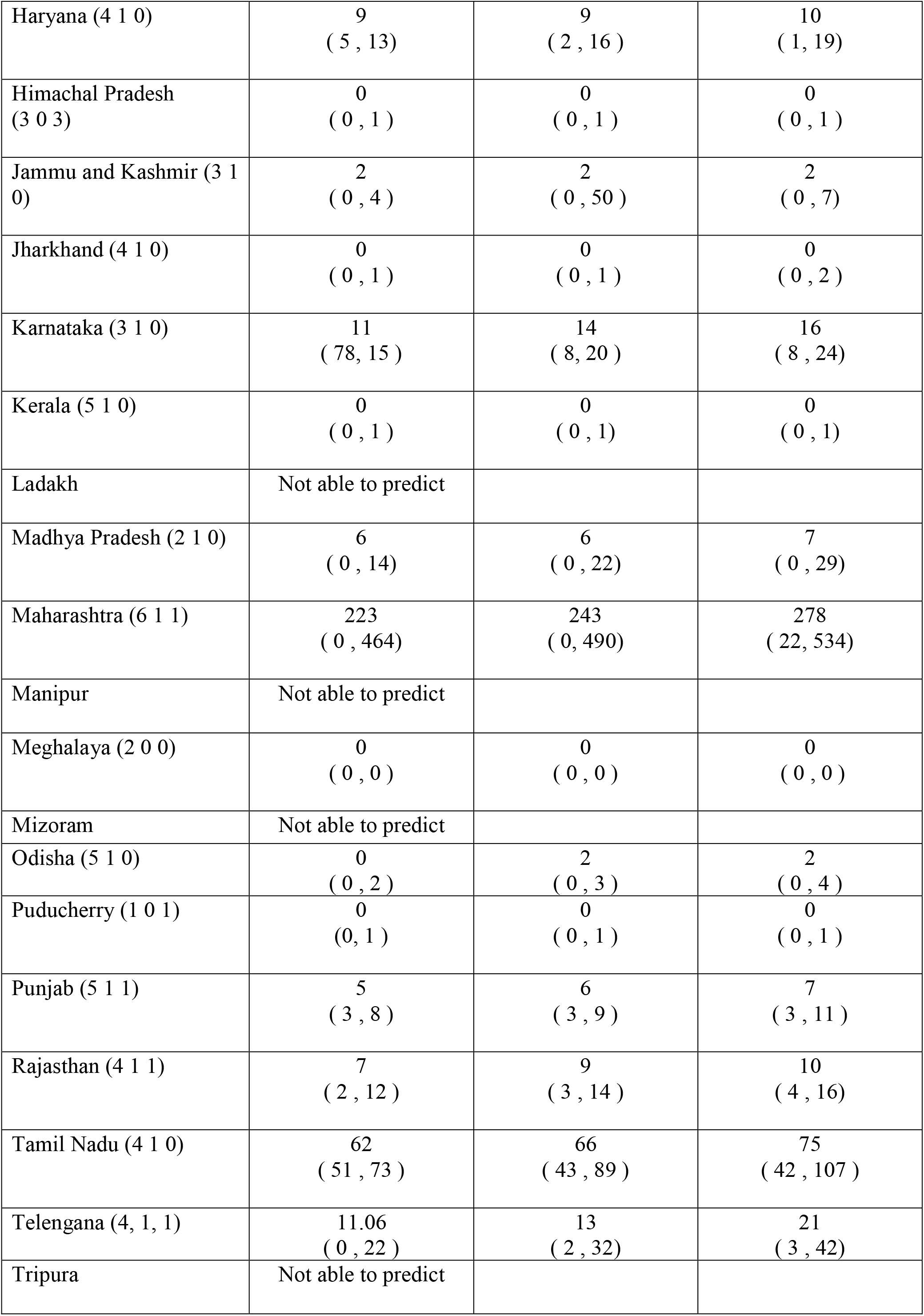
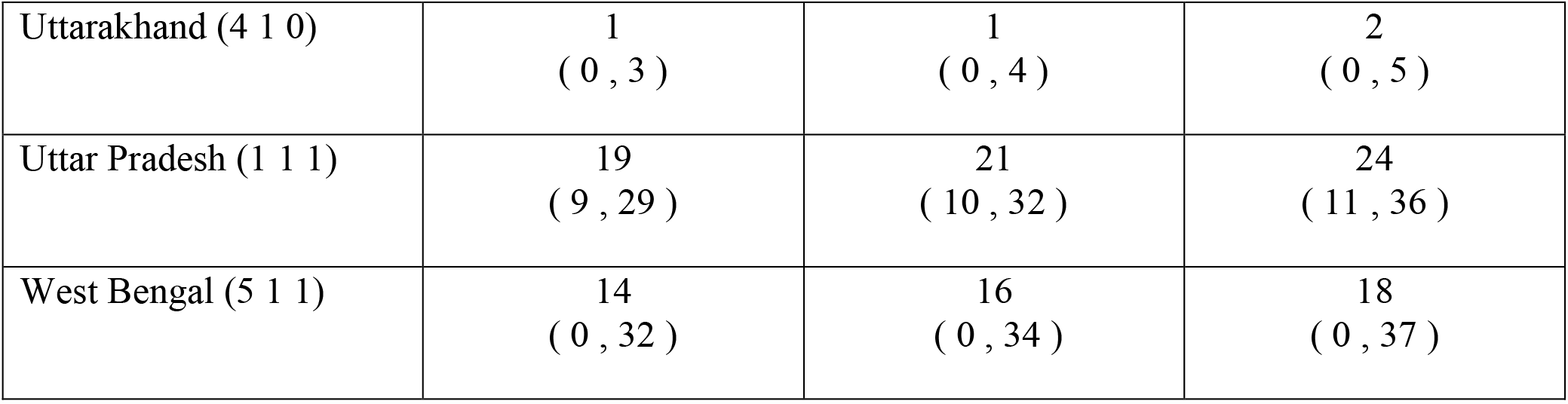
Predicted Daily Fatality for each state and UT with 95% CI

| State & Model with<br>(p, d, q) | 02 July 2020<br>Predicted value<br>(lower limit, upper limit) | 15 July 2020<br>Predicted value<br>(lower limit, upper limit) | 01 August 2020<br>Predicted value<br>(lower limit, upper limit) |
| --- | --- | --- | --- |
| Andaman and Nicobar Islands | Not able to predict |  |  |
| Andhra Pradesh (3 1 0) | 11<br>( 8 , 14 ) | 13<br>( 7 , 18 ) | 15<br>( 7 , 22 ) |
| Arunachal Pradesh<br>(3 1 0) | 0<br>( 0 , 0 ) | 0<br>( 0 , 0 ) | 1<br>( 0 , 2 ) |
| Assam (4 1 0) | 0<br>( 0 , 2 ) | 0<br>( 0 , 2 ) | 1<br>(0, 3 ) |
| Bihar (3 1 1) | 2<br>( 0 , 3 ) | 2<br>(0, 4 ) | 2<br>( 0 , 4 ) |
| Chandigarh<br>(4 1 0) | 0<br>(0, 1 ) | 0<br>( 0 , 1 ) | 0<br>(0, 1 ) |
| Chhattisgarh<br>(5 1 0) | 0<br>( 0 , 1 ) | 0<br>( 0 , 1 ) | 0<br>( 0 , 2 ) |
| Dadar Nagar Haveli | Not able to predict |  |  |
| Delhi (6 1 1) | 67<br>(0, 140 ) | 76<br>(0, 156 ) | 88<br>(8, 168) |
| Goa (2 1 0) | 0<br>( 0 , 1 ) | 1<br>( 0 , 1 ) | 1<br>(0, 2) |
| Gujarat (4 1 1) | 18<br>( 7 , 29 ) | 21<br>( 0 , 43 ) | 24<br>(0, 54) |
| Haryana (4 1 0) | 9<br>( 5 , 13) | 9<br>( 2 , 16 ) | 10<br>( 1 , 19) |
| Himachal Pradesh<br>(3 0 3) | 0<br>( 0 , 1 ) | 0<br>( 0 , 1 ) | 0<br>( 0 , 1 ) |
| Jammu and Kashmir (3 1 0) | 2<br>( 0 , 4 ) | 2<br>( 0 , 50 ) | 2<br>( 0 , 7) |
| Jharkhand (4 1 0) | 0<br>( 0 , 1 ) | 0<br>( 0 , 1 ) | 0<br>( 0 , 2 ) |
| Karnataka (3 1 0) | 11<br>( 78, 15 ) | 14<br>( 8, 20 ) | 16<br>( 8 , 24) |
| Kerala (5 1 0) | 0<br>( 0 , 1 ) | 0<br>( 0 , 1) | 0<br>( 0 , 1) |
| Ladakh | Not able to predict |  |  |
| Madhya Pradesh (2 1 0) | 6<br>( 0 , 14) | 6<br>( 0 , 22) | 7<br>( 0 , 29) |
| Maharashtra (6 1 1) | 223<br>( 0 , 464) | 243<br>( 0 , 490) | 278<br>( 22, 534) |
| Manipur | Not able to predict |  |  |
| Meghalaya (2 0 0) | 0<br>( 0 , 0 ) | 0<br>( 0 , 0 ) | 0<br>( 0 , 0) |
| Mizoram | Not able to predict |  |  |
| Odisha (5 1 0) | 0<br>( 0 , 2 ) | 2<br>( 0 , 3 ) | 2<br>( 0 , 4 ) |
| Puducherry (1 0 1) | 0<br>(0, 1 ) | 0<br>( 0 , 1 ) | 0<br>( 0 , 1 ) |
| Punjab (5 1 1) | 5<br>( 3 , 8 ) | 6<br>( 3 , 9 ) | 7<br>( 3 , 11 ) |
| Rajasthan (4 1 1) | 7<br>( 2 , 12 ) | 9<br>( 3 , 14 ) | 10<br>( 4 , 16) |
| Tamil Nadu (4 1 0) | 62<br>( 51 , 73 ) | 66<br>( 43 , 89 ) | 75<br>( 42 , 107 ) |
| Telangana (4, 1, 1) | 11.06<br>( 0 , 22 ) | 13<br>( 2 , 32) | 21<br>( 3 , 42) |
| Tripura | Not able to predict |  |  |
| Uttarakhand (4 1 0) | 1<br>( 0 , 3 ) | 1<br>( 0 , 4 ) | 2<br>( 0 , 5 ) |
| Uttar Pradesh (1 1 1) | 19<br>( 9 , 29 ) | 21<br>( 10 , 32 ) | 24<br>( 11 , 36 ) |
| West Bengal (5 1 1) | 14<br>( 0 , 32 ) | 16<br>( 0 , 34 ) | 18<br>( 0 , 37 ) |

**Table 3:** Ventilator quantity requirement state wise

| States | Fatality Rate | Ventilator Requirements<br>as on 01 August 2020 |
| --- | --- | --- |
| Andaman and Nicobar Islands | 0.0050 | 81 |
| Andhra Pradesh | 0.0008 | 808 |
| Arunachal Pradesh | 0.0004 | 4 |
| Assam | 0.0005 | 233 |
| Bihar | 0.0040 | 1945 |
| Chandigarh | 0.0002 | 4 |
| Chhattisgarh | 0.0001 | 14 |
| Dadar Nagar Haveli | - | 3 |
| Delhi | 0.0021 | 9794 |
| Goa | 0.0090 | 120 |
| Gujarat | 0.0035 | 5104 |
| Haryana | 0.0006 | 473 |
| Himachal Pradesh | 0.0029 | 132 |
| Jammu and Kashmir | 0.0011 | 399 |
| Jharkhand | 0.0013 | 153 |
| Karnataka | 0.0022 | 2856 |
| Kerala | 0.0001 | 34 |
| Ladakh | - | 3 |
| Madhya Pradesh | 0.0028 | 1624 |
| Maharashtra | 0.0028 | 26114 |
| Manipur | - | 3 |
| Meghalaya | 0.0014 | 3 |
| Mizoram | - | 3 |
| Odisha | 0.0003 | 119 |
| Puducherry | 0.0009 | 25 |
| Punjab | 0.0052 | 1296 |
| Rajasthan | 0.0007 | 526 |
| Tamil Nadu | 0.0007 | 3951 |
| Telangana | 0.0016 | 317 |
| Tripura | - | 3 |
| Uttarakhand | 0.0004 | 59 |
| Uttar Pradesh | 0.0009 | 1033 |
| West Bengal | 0.0033 | 3228 |

Proposed methodology for ventilator quantity requirement is calculated from the fatality rate. Fatality rate is calculated as the ratio of daily fatality and confirmed case. Mean of the fatality rate is calculated along with predicted confirmed upper limit value is used for finding the ventilator requirement. Andaman and Nicobar Islands, Dadar Nagar Haveli, Ladakh, Manipur, Mizoram and Tripura state prediction for fatality with ARIMA model is not possible because of the property of the data as discussed earlier (death case). Daily fatality for these states is zero for all reported dates. These states are assigned three as ventilator requirement which is the minimum value from all other states because these states are having zero as the fatality. Total number of ventilators required in India as on 01 August 2020 is 60,504.

## 5. Conclusion

COVID 19 has been declared as a pandemic by WHO and is currently a major global threat. This study have conducted to find the cumulative confirmed cases, fatality rate and ventilator requirement for Indian states in near future. This study helps the administrators in each state to take decision regarding the induction of ventilators in each state and this will help the health workers to prepare for future. ARIMA model is selected for predicting the future confirmed cases and fatality in Indian states. As per the model, confirmed cases are increasing in the future and the requirement for ventilators are also increasing. But the actual numbers can go high due to the following factors such as social distancing, testing capacity, population density, age structure, societal collectivism, repatriation flights from different countries and movement of informal workers.

## Data Availability

Data pertaining to this work is available publicly available in https://www.kaggle.com/

https://www.kaggle.com/sudalairajkumar/covid19-in-india?select=covid_19_india.csv

## 6. Acknowledgment

We are most indebted to Mrs. Manimozhi Theodore, Outstanding Scientist & Director of Defence Bio-Engineering and Electro Medical Laboratory (DEBEL), Bangalore, India for her initiative and invaluable constant support which enabled us to continue and complete this work. We would like to thank Technical Coordination Group and Human Resource Department of DEBEL for sincerely helping us in publishing this paper.

